# The TBVaxRepository: A living database of projects supporting the preparedness for adult and adolescent TB vaccine rollout

**DOI:** 10.64898/2026.05.06.26352615

**Authors:** Joeri Sumina Buis, Andrew D. Kerkhoff, Christiaan Mulder, Degu Jerene

## Abstract

**Background:** Tuberculosis (TB) continues to cause substantial morbidity and mortality, with adults and adolescents carrying the largest burden of disease. Multiple promising novel vaccine candidates are in clinical trials, and their eventual impact will depend on effective implementation strategies. Information on TB vaccine preparedness efforts that could inform coordination remains fragmented.

**Methods:** We developed the first living and interactive online repository (https://tbvaxrepository.org/) collating completed, ongoing, and planned adult and adolescent TB vaccine preparedness initiatives. Data were obtained through a prior scoping review, direct stakeholder engagement, international conferences, and open calls via social media and partner networks between March 2023-November 2024. Projects were categorized using the World Health Organization’s (WHO) framework for TB vaccine preparedness across three thematic areas: availability, accessibility, and acceptability.

**Findings:** By December 2024, the repository included 90 projects from 119 countries. Most projects focused on health- (47%) and economic modelling (21%), demand and acceptability studies (19%) or implementation feasibility (14%). Most of the projects were situated in India (n=36), South Africa (n=34), China (n=19), Indonesia, (n=17), Kenya (n=17), Brazil (n=14), and Pakistan (n=14). Few initiatives targeted key populations such as people living with HIV, pregnant or lactating individuals, or socially marginalized and occupational high-risk groups. Research on communication strategies for facilitating uptake as part of rollout were absent.

**Conclusions:** The repository reveals both progress and gaps in global TB vaccine preparedness across WHO’s three thematic areas, with particular attention to geographic coverage, and the inclusion of key populations. As novel vaccines for adults and adolescents approach potential licensure, coordinated and inclusive preparedness efforts will be critical to ensure equitable and effective rollout. This repository offers a transparent platform to strengthen collaboration, reduce duplication, and guide strategic planning in a historically underfunded field.

## Introduction

Tuberculosis (TB), a disease that has afflicted humanity for over 70,000 years, still claimed a life every 25 seconds in 2024, despite significant progress in combating it (1,2). A novel tuberculosis (TB) vaccine for adults and adolescents is regarded as a necessary breakthrough to accelerate the reduction in TB-related morbidity and mortality (3). To date, only one licensed TB vaccine exists -the Bacillus Calmette Guerin (BCG) vaccine. While BCG provides protection against meningeal, disseminated, and pulmonary TB in children, it offers little to no protection for adults and adolescents, the group with the largest TB burden worldwide (4–6). Encouragingly, multiple TB vaccine candidates targeting these populations have progressed along the clinical pipeline, with some, showing promising preliminary results (7).

Currently, 17 TB vaccine candidates include adolescents and/or adults in their clinical trials, with 3 in active phase 3 trials or starting soon (8,9). These vaccines are being evaluated for either the prevention of disease (PoD) or prevention of infection (PoI) and so far, one candidate has demonstrated promising vaccine efficacy (VE) results. A M72/AS01e phase 2b trial found a VE of 49.7% against the development of TB disease over a three year follow-up period (90% confidence interval [CI] = 12.1 to 71.2 and 95% CI = 2.1 to 74.2) (10). The results of phase 3 clinical trial results for M72/AS01e, enrolling 20,000 adult and adolescent participants across five countries, are expected as early as 2028 (11). While stakeholders were hopeful about the BCG revaccination candidate, the most recent BCG revaccination trial was discontinued after results from its 2b trial showed no protective effect from the BCG revaccination when comparing participants who received the BCG compared to the placebo group (risk ratio of 1.09 [95% CI 0.67–1.77]; p=0.791) (12).

While the development of new TB vaccines is foundational, it is vaccinations—not vaccines alone—that save lives. Even if one of these candidates proves efficacious against TB disease in adults and adolescents, its impact will depend on effective implementation strategies (13,14). With few vaccine programs intended for adults and adolescents, existing immunization platforms and strategies for these groups are limited. In response to this, the World Health Organization (WHO) created a global framework to “guide countries and global stakeholders in preparing for the introduction and coverage scale-up of new TB vaccines for adolescent and adult populations”(14). This framework aims to support countries and global stakeholders in scaling up coverage of novel TB vaccines by addressing *availability*, *accessibility*, and *acceptability* with the aim of preparing health systems and communities for successful vaccine rollout.

Our prior scoping review identified significant gaps in knowledge across multiple domains (Epidemiological impact, economic impact, acceptability, and implementation feasibility) (15).

Encouragingly, driven by the prospect of a vaccine that could be available in the next 5 years, several activities to prepare for effective rollout are being implemented or planned. However, lack of reliable information on completed, ongoing and planned initiatives can cause duplication of efforts leading to wastage of meagre resources or under coverage of preparedness activities in certain countries and settings. We developed a living and interactive online repository capturing completed, ongoing, and future planned adult and adolescent TB vaccine preparedness initiatives with the aim of enhancing global collaboration, guide strategic planning, and identify critical gaps in preparedness. This article outlines the methodology and content of the repository, highlighting how it can drive global coordination, strategic planning, and facilitate an improved rollout of novel TB vaccines for adolescents and adults.

## Methods

### Project identification

We used five approaches to collect and triangulate data on completed, ongoing, and planned projects. First, we identified completed research based on our previously conducted scoping review on TB vaccine preparedness (15). Then, we requested updates on completed, ongoing, and planned projects through e-mail using a newly-built stakeholder base (details below). Additionally, stakeholders were asked to share the request for updates with other relevant stakeholders. We attended and spoke to stakeholders regarding projects at previously identified relevant international meetings and global conferences (the WHO meetings on TB vaccine preparedness, IAVI’s TB Vax ARM meetings, the Global Forum on TB Vaccines (GFTBV), and the Union conference) (16–18).

Lastly, we requested updates through social media posts (LinkedIn), newsletters, and websites of collaborating organizations to share data through the repository website once the repository was launched.

### Building a stakeholder base

We built a stakeholder base consisting of stakeholders involved in TB vaccine preparedness work. Stakeholders included researchers and policy makers within the TB vaccine preparedness field. We identified stakeholders through connections of the Supporting, Mobilizing, and Accelerating Research for Tuberculosis Elimination (SMART4TB) consortium (19). Additional stakeholders were identified via WHO’s meetings on TB vaccine preparedness, International AIDS Vaccine Initiative’s (IAVI) TB Vaccine Advocacy Roadmap (TBVAX ARM) meetings, the Global Forum for TB Vaccines (GFTBV), Union conference, and snowballing when contacting the stakeholders (16–18).

### Data collection and the repository

We collected data using an Excel sheet with the respective project details to be filled in (see table S.1). The excel sheet was available on the repository website and shared with stakeholders when contacted for updates. We reached out twice to the stakeholder base for updates between March 2023 and November 2024. Additionally, we updated the repository when new information was shared or identified.

We categorized projects using themes and subthemes derived from the WHO’s global framework for TB vaccine preparedness (14). Projects could be categorized under multiple (sub)-themes if applicable:

- *Availability*: epidemiological impact, economic impact and cost-effectiveness, policy and frameworks, and procurement,
- *Accessibility*: implementation strategies and feasibility, health system readiness, and financing strategies,
- *Acceptability*: acceptability and demand, community engagement, and communication strategies.

Additional details captured included countries, target groups, vaccine candidate, type of protection of the vaccine (PoI, PoD, Prevention of Recurrence (PoR)), type of project (policy and frameworks, research, advocacy, other) and type of research (modelling, quantitative, qualitative, mixed-method, other), timeline, partners involved, existing resources/tools, and a contact person.

Feedback rounds held with SMART4TB’s Community Advisory Board (CAB), the WHO, and stakeholders included in the stakeholder base ensured the alignment with the needs of the field and accessibility of the tool.

### Data analysis

We summarized data descriptively using StataCorp Standard Edition (SE) 17 and reported results according to the WHO global framework for TB vaccine preparedness (sub)-themes and by country (14,21). Using the geographic information system QGIS (22), we mapped the number of TB vaccine preparedness projects against country-level TB, TB/HIV, and rifampicin resistant (RR)/multi drug resistant (MDR) TB burden indicators. Specifically, the number of project was mapped for each of the following WHO indicators: 1. estimated total incident TB cases, 2. TB incidence per 100,000 population, 3. total TB/HIV cases, 4. TB/HIV as a percentage of total TB cases, 5. Total RR/MDR TB cases, and 6. RR/MDR TB as a percentage of total TB cases (22,23). We ranked and compared WHO’s high priority countries (TB, TB/HIV, and/or RR/MDR TB) with their respective ranking by number of TB vaccine preparedness projects (24).

## Results

### General characteristics

We officially launched the repository website at the Union Conference in November 2024 and last updated the repository end of December 2024 (https://tbvaxrepository.org/; Figure 1). By the end of 2024, the repository included 90 projects, and we built a stakeholder base consisting of 28 organizations, totalling 55 experts (table 1). We identified 65 research projects, 10 policy/frameworks, 6 advocacy projects, and 7 other types of projects (table S.2).

**Figure 1.**
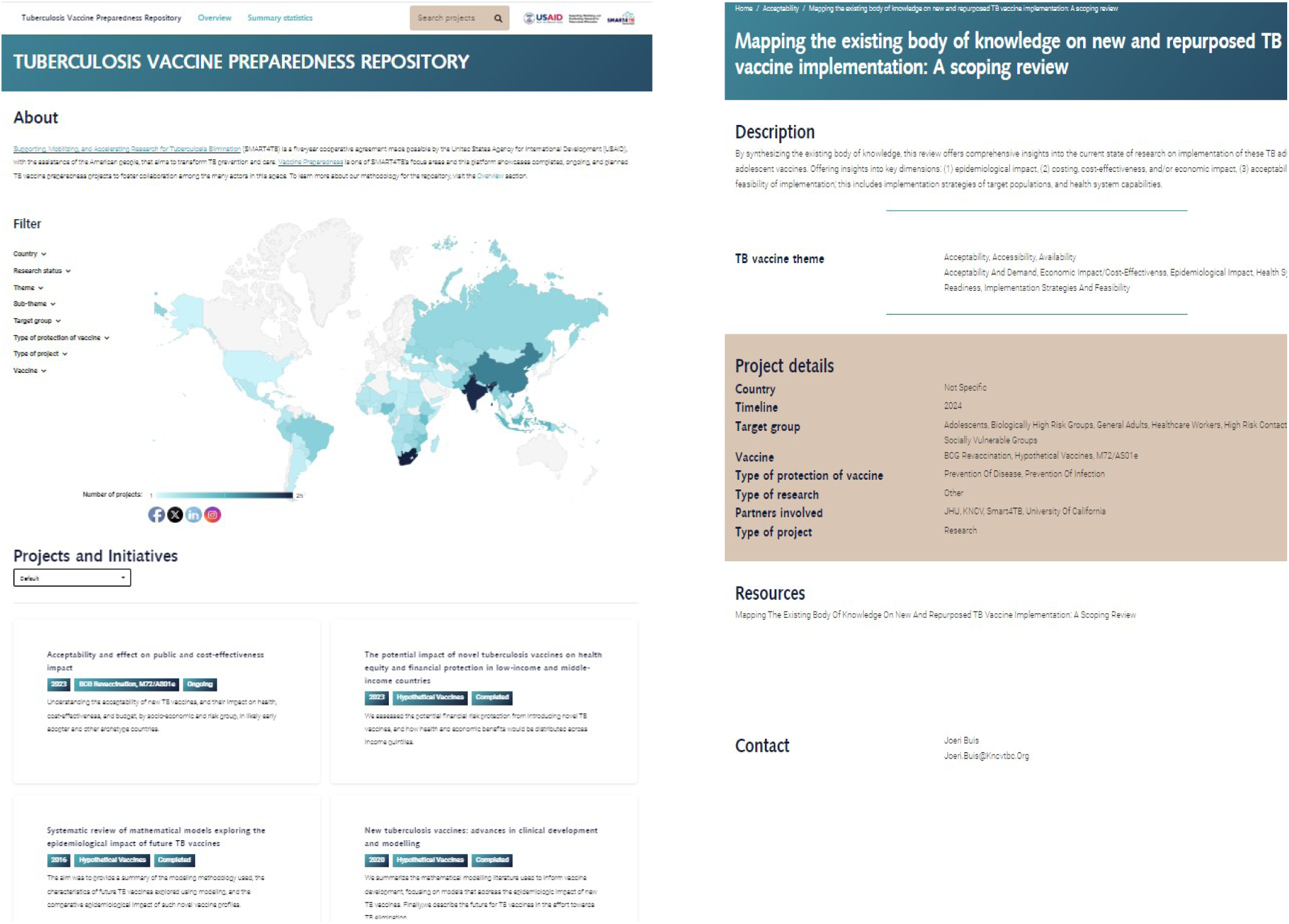
On the left: the main page of the TB vaccine preparedness repository website. On the right: Example of a project page of the TB vaccine preparedness repository website. The website holds details of the identified completed, ongoing, and future planned adult and adolescent TB vaccine preparedness projects as of December 2024.

**Table 1.** Overview of identified stakeholder experts and projects per method. This excludes previously identified experts/projects by another method.

| **Approaches to collect data** | **Stakeholders identified** | **Completed projects** | **Ongoing projects** | **Planned projects** | **Total projects identified** |
| --- | --- | --- | --- | --- | --- |
| Scoping review | - | 25 | - | - | 25 |
| Stakeholder base and snowballing | 33 | 20 | 11 | 7 | 38 |
| International meetings and conferences | 22 | - | 27 | - | 27 |
| Social media | 0 | - | - | - | 0 |
| Website of the repository | 0 | - | - | - | 0 |
| Total projects |  | 45 | 38 | 7 | 90 |

#### Timeline and thematic approach of projects

In the repository, the initiation date of the projects spanned between 2011 to 2024. Twenty projects (22%) did not have a clearly specified project initiation date. As shown in figure 2, the number of projects gradually increased until 2022, followed by a large increase in 2023 and 2024, when more than 10 projects were started each year. Most projects in the first 10 years focused on *availability*. However, starting from 2022, more newly initiated projects focused on *accessibility* and/or *availability* (figure 2). Of the 90 projects, 67 (74%) included aspects of availability, 19 (21%) of acceptability, and 16 (17%) of accessibility. Among projects focusing on *availability*, most modelled health impacts (63%), followed by economic impact and cost-effectiveness (28%), and policy/frameworks (25%). Among projects including *accessibility* aspects, most assessed implementation strategies (81%), followed by financing strategies (25%), procurement (25%), and health system preparedness (19%). Among projects covering *acceptability*, most aimed to understand demand/acceptability (89%), followed by community engagement (32%). No projects were identified that included activities focusing on designing communication strategies as part of vaccine implementation efforts (table S.3 and table S.4). Figure 3 shows an overview of completed (n=45), ongoing (n=38), and planned (n=7) projects across the world. Projects with a start date after 2024 are planned in Nigeria, South Africa (SA), Myanmar, Vietnam, Indonesia, China, India, DRC, Brazil, Ethiopia, Kenya, Pakistan, Bangladesh, and Tajikistan - all countries that also had ongoing and completed projects.

**Figure 2.**
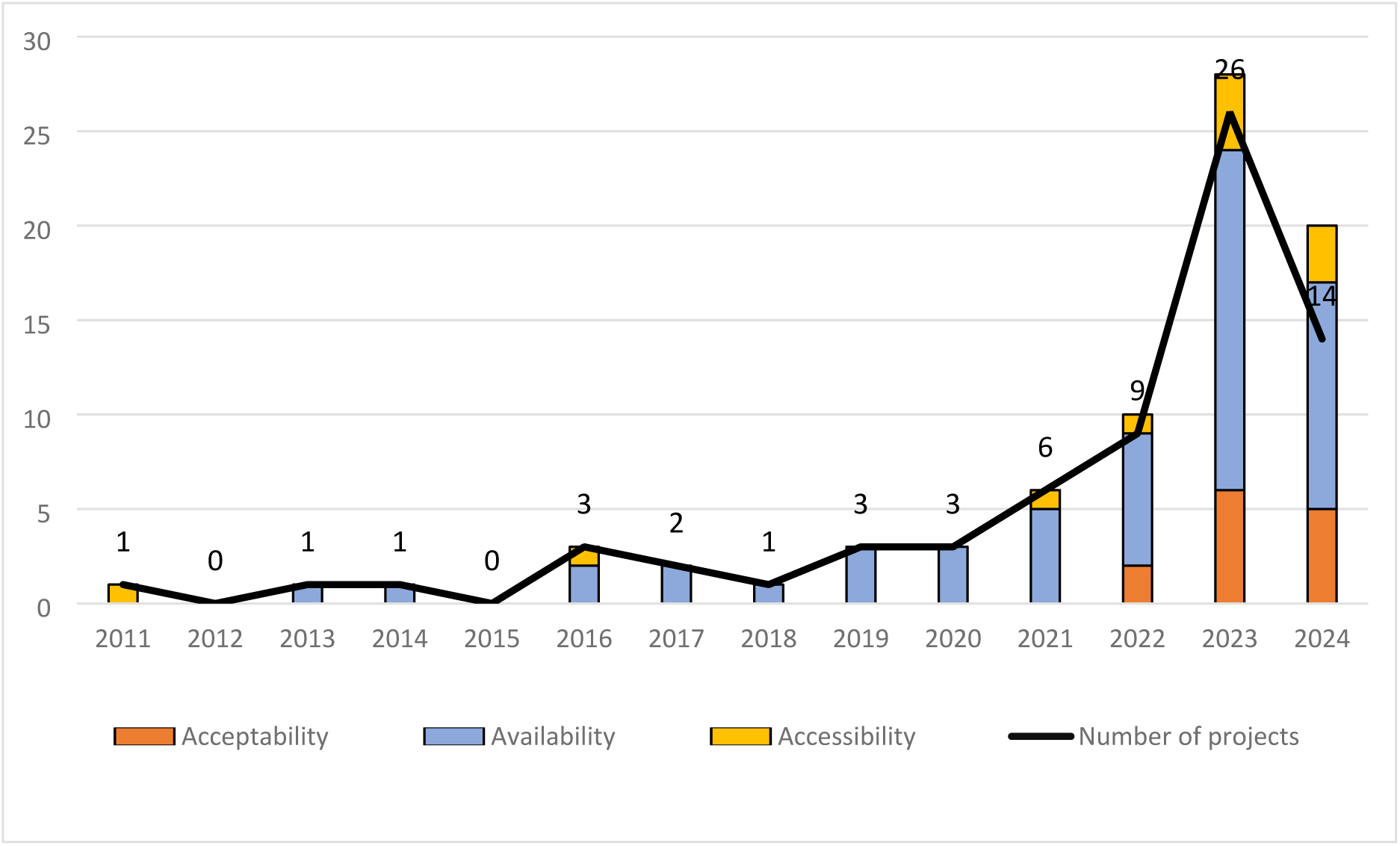
Timeline of the number of newly initiated and ongoing projects per year and WHO’s global framework for TB vaccine preparedness theme “acceptability”, “availability”, and “accessibility”. The black line shows the number of projects per year included in the TB Vaccine Preparedness repository. The bars indicate the themes covered by the projects per year. Of the 90 projects included in the repository, 20 projects (22%) did not have information regarding initiation date. Note: a single project may cover multiple themes.

**Figure 3.**
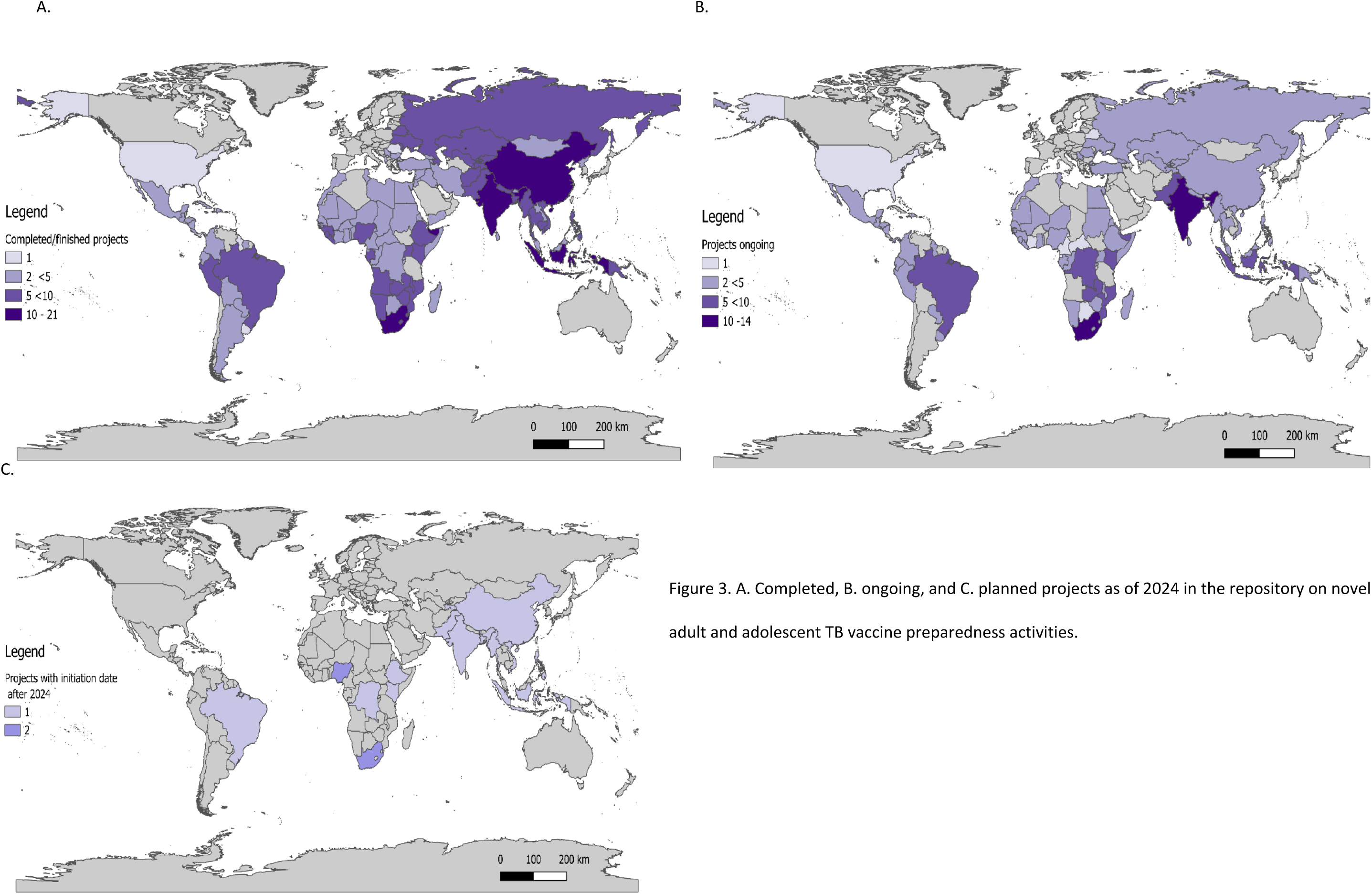
A. Completed, B. ongoing, and C. planned projects as of 2024 in the repository on novel adult and adolescent TB vaccine preparedness activities.

### Country representation

#### General

Of the 215 countries/territories listed by the WHO, 119 had at least one project regarding novel adult and adolescent TB vaccine preparedness (see table S.4). In all 119 countries, projects included aspects of epidemiological impact, and economic impact and cost-effectiveness. In 14 countries, projects included procurement aspects; in another 14, projects included aspects of financing strategies. Projects focusing on acceptability and demand spanned 49 countries, implementation strategies 51 countries, health system preparedness 47 countries, and community engagement 1 country. SA was the only country including all sub-themes except communication strategies. Most projects were concentrated in India (n=36) and SA (n=34), followed by China (n=19), Indonesia, (n=17), Kenya (n=17), Brazil (n=14), and Pakistan (n=14) (see table S.5).

#### TB burden

The figures visualizing the number of projects against WHO’s estimated TB burden show a wide distribution globally, with a concentration in countries with the highest total estimated TB cases (see figure 4). All WHO high burden priority countries had at least 5 projects on novel adult and adolescent TB vaccine preparedness, except for the Democratic People’s Republic of Korea (n=2).

**Figure 4.**
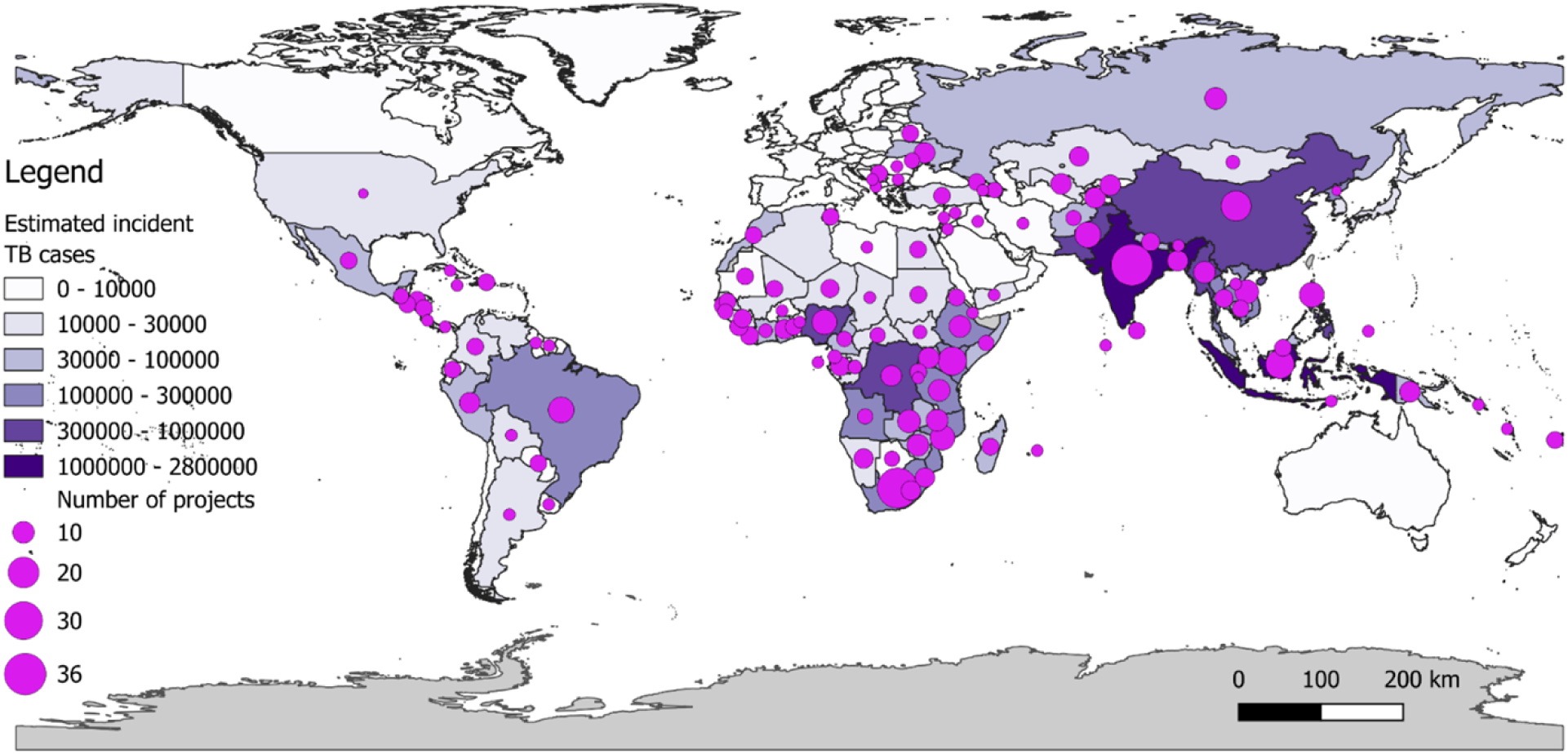
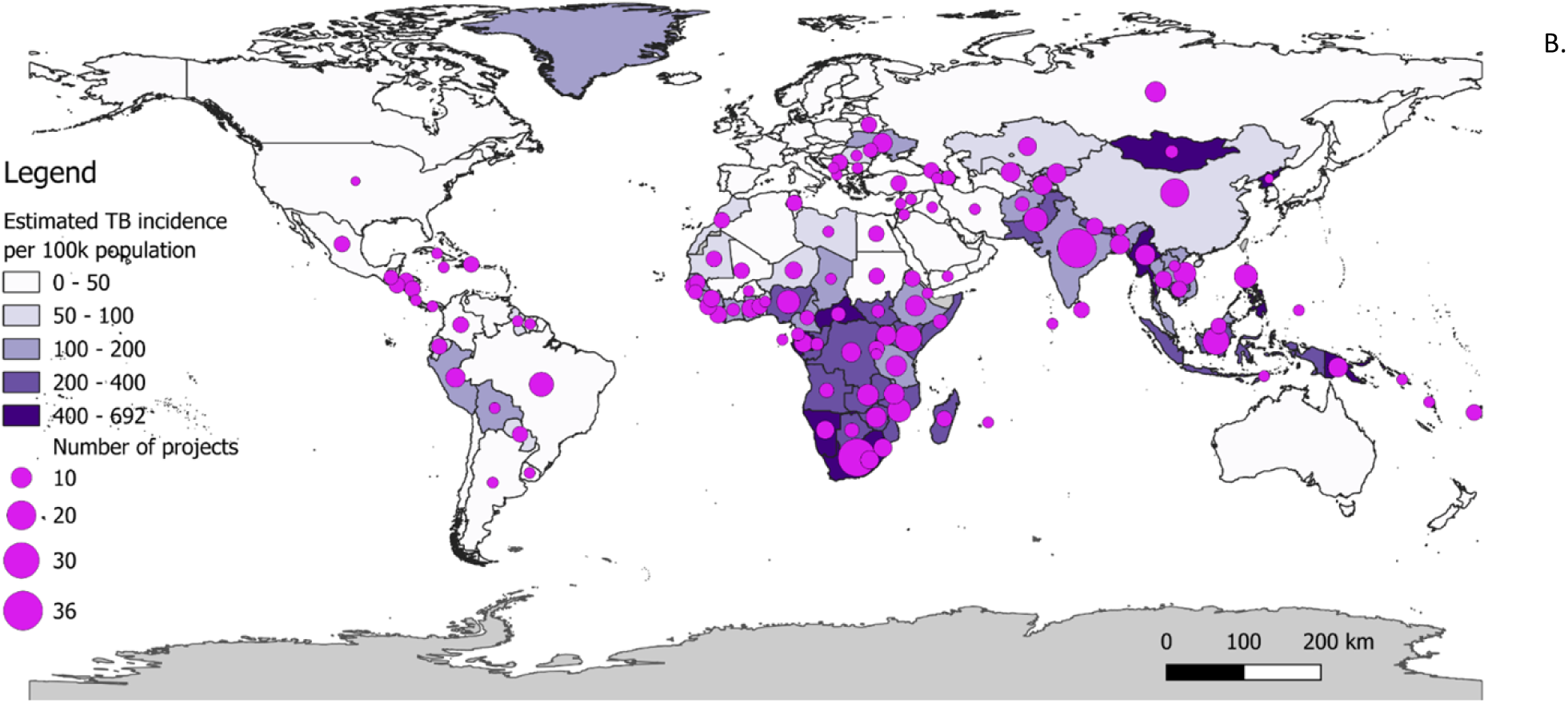
A. World map with the number of adult and adolescent TB vaccine preparedness projects/initiatives and the estimated incident TB cases in 2023, B. estimated TB incidence per 100k population (25).

Priority countries ranking high in total incident TB cases but relatively lower in number of projects were Angola, Bangladesh, Cameroon, CAR, Congo, DRC, Democratic People’s Republic of Korea (North Korea), Mongolia, Somalia, and Thailand. Priority countries that ranked high according to their total TB incidence per 100,000 but relatively lower on number of projects were Angola, Botswana, Cameroon, CAR, Congo, North Korea, Eswatini, Gabon, Guinea-Bissau, Lesotho, Liberia, Mongolia, Namibia, Papua new guinea, Sierra Leone, and Somalia (see S.5).

#### TB/HIV burden

The figures visualizing the number of projects against WHO’s estimated TB/HIV burden show that most projects are conducted in the countries with the highest estimated total TB/HIV cases (figure S.1). All WHO high burden priority countries had at least 2 projects on novel adult and adolescent TB vaccine preparedness, except for North Korea. Priority countries that ranked high according to their TB/HIV total cases and had a relatively lower ranking in the number of projects were Angola, Botswana, Cameroon, CAR, Congo, DRC, North Korea, Guinea-Bissau, Thailand, and Uganda. Priority countries that ranked high according to TB/HIV cases as a percentage of total estimated TB cases and had relatively lower ranking in number of the projects were Botswana, Cameroon, CAR, Congo, North Korea, Eswatini, Gabon, Guinea-Bissau, Lesotho, Malawi, Moldova, Uganda, and Zimbabwe (see S.5).

#### RR/MDR TB burden

The figures visualizing the number of projects and the WHO’s estimated RR/MDR TB burden showed less alignment with respective burden (figure S.2). All WHO high burden priority countries had at least 2 projects on novel adult and adolescent TB vaccine preparedness, except for North Korea.

Priority countries that ranked high according to their RR/MDR total TB cases but had a relatively lower ranking in the number of projects were Angola, Azerbaijan, Belarus, Cameroon North Korea, Kazakhstan, Mongolia, Nepal, Moldova, Russia, Somalia, Thailand, and Ukraine. Priority countries that ranked high according to the RR/MDR cases as percentage of total estimated TB cases but had a relatively lower ranking in number of projects were Angola, Azerbaijan, Belarus, Botswana, CAR, Congo, North Korea, Kazakhstan, Mongolia, Moldova, Russia, Tajikistan, Ukraine, and Uzbekistan (see S.5).

### Population groups and vaccine characteristics

Most completed, ongoing, and future planned projects include adults and/or adolescents as target groups (n=45 and 46) (figure 5). Only a small number of projects include specific risk groups such as people living with HIV (PLHIV), people with social or biological risk factors, healthcare workers, high risk contacts or occupations, people with drug resistant TB, elderly, and pregnant/breastfeeding people. Most projects assumed a PoD vaccine and used a hypothetical vaccine (rather than a specific candidate), while M72/AS01e was the specific vaccine candidate most commonly assessed (table S.4).

**Figure 5.**
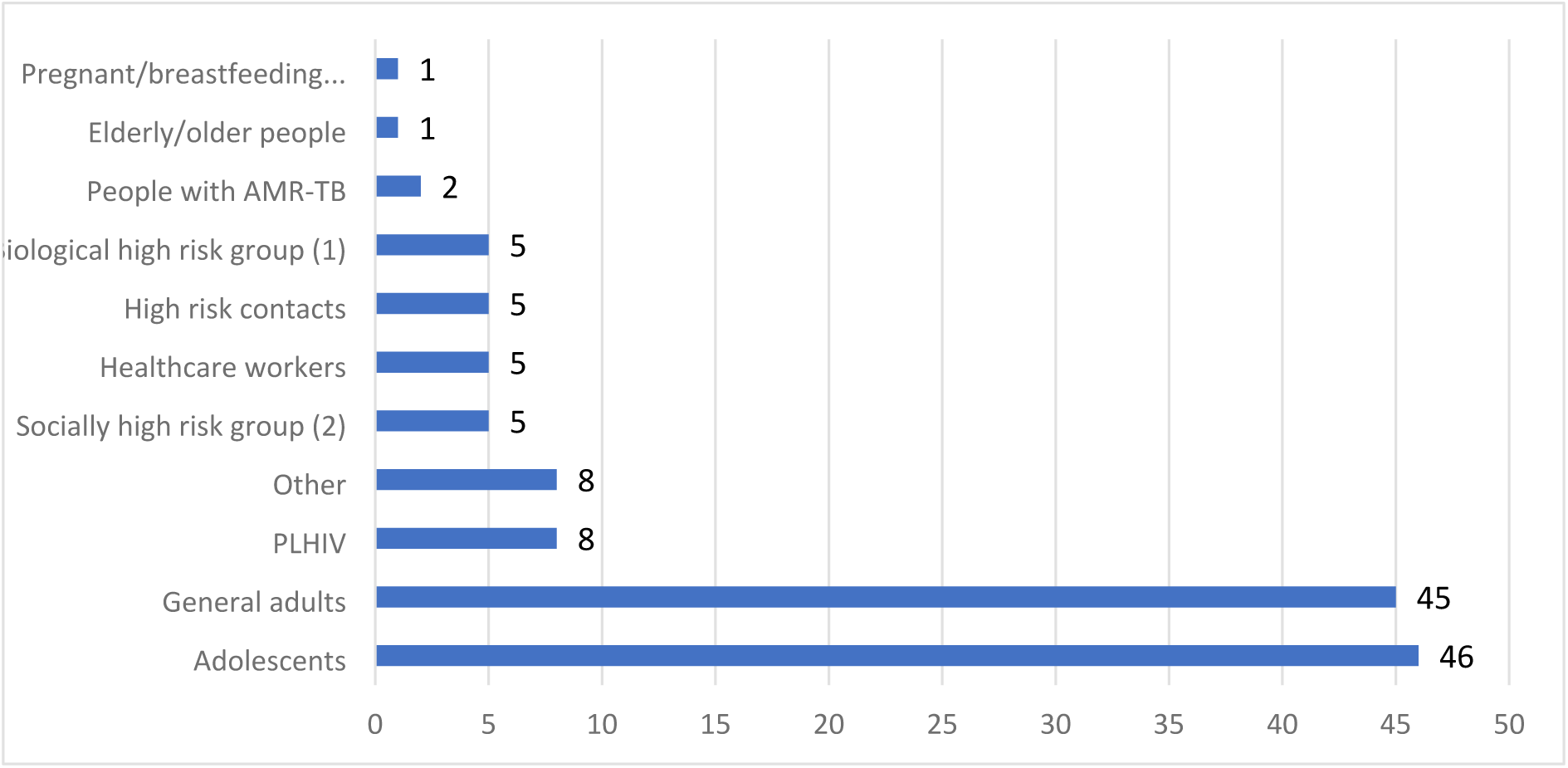
Overview of the number of projects that include pregnant/breastfeeding people, elderly/older people, people with rifampicin resistant/multidrug resistant (RR/MDR) -TB, biological high risk, high risk contacts of people with TB, healthcare workers, socially vulnerable groups, PLHIV, general adults and adolescents as represented in the projects of the TB vaccine preparedness repository. *(1) biological high risk groups = immunocompromised, people with diabetes, (2) socially vulnerable groups = people with alcohol/drug abuse, people deprived of liberty, homeless people.

## Discussion

We created a living and interactive online repository holding the completed, ongoing, and future planned adult and adolescent TB vaccine preparedness projects to enhance global collaboration, support strategic planning, and identify critical gaps in preparedness. The online repository visualized the contents in an informative, accessible, and user-friendly manner increasing its use for various interested users (20). By end of 2024, the repository included 90 projects covering 119 countries. The focus of adult and adolescent TB vaccine preparedness activities has largely focused on understanding health and economic impacts of a novel TB vaccine. In recent years, the focus has shifted to also include aspects regarding accessibility and availability. Most TB vaccine preparedness projects are in countries with the highest TB burden according to total TB cases, yet there are various WHO priority countries with limited projects and little focus on preparedness beyond public health and economic impacts.

Repositories can facilitate streamlining and guiding of global efforts. A recent example of such a global coordination platform is the COVID-19 tracker and its accompanying living mapping review (26,27). The tracker, a live database of all funded COVID-19 research projects facilitated funding coordination and informed decision-making. Such tools underscore the value of centralized, real-time platforms for tracking and guiding projects (27). The findings of our repository highlight a need to ensure equitable country representation in adult and adolescent TB vaccine preparedness activities. SA and India have most projects so far, and both are also considered early adopters; they have ongoing TB vaccine clinical trials, a high TB burden and especially in SA, various in-country activities(28–31). More efforts are needed to initiate preparedness activities in other priority countries. As described in EDCTP’s Global Roadmap for Research and Development for TB Vaccines important elements to ensure public health impact post approval is engagement with decision makers, understanding the TB epidemic and the potential health and economic impact of a novel TB vaccine, ensure post licensure monitoring & evaluation, understand and increase preparedness of the health system, and lastly understand barriers and enablers of acceptability and uptake of a potential novel TB vaccine (32). More projects on accessibility and acceptability are being started however, these activities should be widespread. Identifying which vaccine preparedness findings are transferable across epidemiological, health system, and socio-political contexts can inform strategic prioritization of limited resources. Further, widening the focus beyond general adult and adolescent community members to target groups as well as understudied topics such as community engagement and communication strategies are urgently needed.

Key populations such PLHIV and pregnant and lactating people are at an increased risk of developing and having more severe TB disease, making the need for a novel adult and adolescent TB vaccine evident. Meanwhile, these groups often have special considerations when included in clinical trials. Previous unanticipated harm, legal liabilities and insufficient market interest are preventing the inclusion of these groups in current vaccine trials (33). Increasingly however, stakeholders are calling for key populations to be included in trials and research. For example, the WHO recently launched a call to action to promote “*the protection of pregnant and lactating people **through** research and not **from** research*”(34). The exclusion of these groups in current vaccine trials could partially explain the exclusion of these groups in preparedness research. Preparedness research however, could further advocate for the inclusion of key groups by demonstrating potential demand, preferred implementation strategies, and potential impact. More advocacy and research centring the voices of those most affected by TB is necessary to ensure acceptable and effective implementation strategies (35).

Global and local efforts are needed to ensure optimal implementation. The WHO has created various guidance documents and has been conducting workshops in Indonesia and SA to understand what is necessary for effective out roll (16,36). Additionally, there are various advocacy and community efforts such as the TB Vax Arm (37) and the TB CAB (38) to ensure engagement and representation. One current gap the repository highlights is the lack of projects on communication strategies. How do we reach target populations efficiently and effectively by designing and disseminating resonant and trustworthy messages that promote vaccine confidence?

Strengths of this study include the creation of the first-of-its kind global, freely accessible repository centralizing global preparedness activities for implementation of a novel adult and adolescent TB vaccine (26). We used diverse engagement methods, including attending international meetings, building a standard stakeholder base, raising awareness through social media to systematically identify global activities. Especially attending meetings and conferences helped identify ongoing, and future planned activities while the previously conducted scoping review helped identify completed (research) projects. Limitations of the repository include the loss of funding that has not (as of date of publication) been replaced to keep the repository updated and adjusted to the needs of the field due to the executive decisions made by the US government in early 2025(39). Additionally, activities especially in Russia or China may be underrepresented given the lack of stakeholder representation in the stakeholder base. Further, this repository does not capture the individual efforts that governments may be undertaking or planning and are not in the public domain nor does it evaluate the preparedness of the country to effectively rollout a novel TB vaccine for adults and adolescents. The current repository was dependent on manual requesting of data and imputing of data into the repository. To minimize resources and ensure the sustainability of this tool, automatic uploading and checks by individual stakeholders with a live review similar to the COVID live review, would benefit the sustainability of the tool.

## Conclusion

There is an urgent need for equitable and streamlined coordination of TB vaccine preparedness efforts globally to support a successful rollout, as soon as the next 5 years. This study and the resultant TB vaccine preparedness repository contribute to that by creating a transparent platform which holds completed, ongoing, and future planned TB vaccine activities. The repository can stimulate collaboration between stakeholders, provide tools for advocacy purposes, and highlights gaps in knowledge and activities - with a notable gap in community demand and acceptance research. Keeping the repository updated and responsive to the evolving needs in the field is paramount to a coordinated and efficient approach to preparing for the successful implementation of a novel adult and adolescent TB vaccine.

## Data Availability

The dataset can be downloaded from the website of the repository (https://tbvaxrepository.org/).

## Acknowledgement

This research was made possible by the support of the U.S. Government through the SMART4TB consortium under cooperative agreement number 7200AA22CA00005. The contents are the sole responsibility of the authors and do not necessarily reflect the views of the United States Government, consortium collaborators or members.

We would also like to thank the following people: We sincerely thank Puck T. Pelzer for her contributions in the initial phases of the conceptualisation of the repository, and Rupali J. Limaye, Arman Majidulla, Kirthini Muralidharan, and Michelle M. Gill for their input and contributions during all phases of the project.

---

**Figure S.1.**
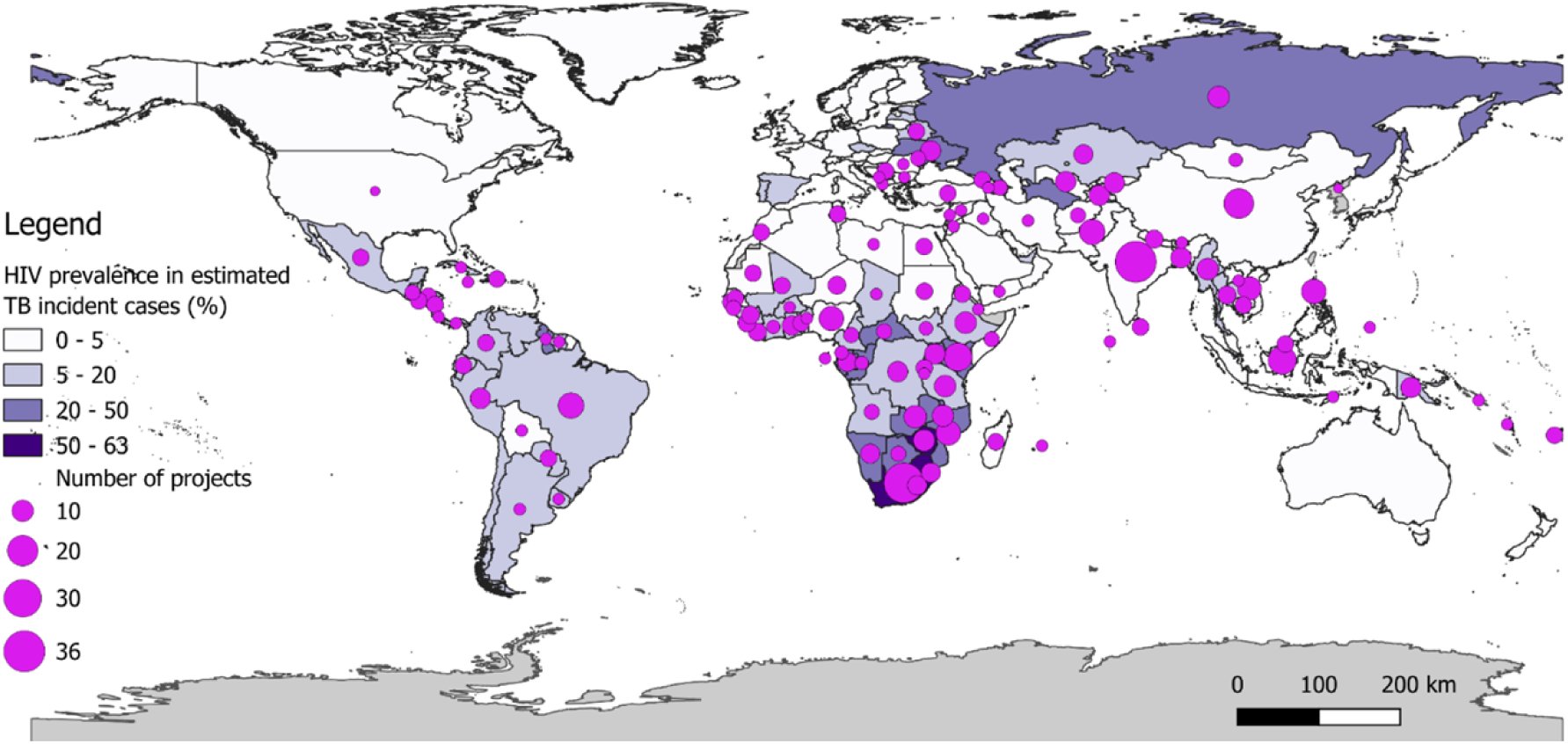
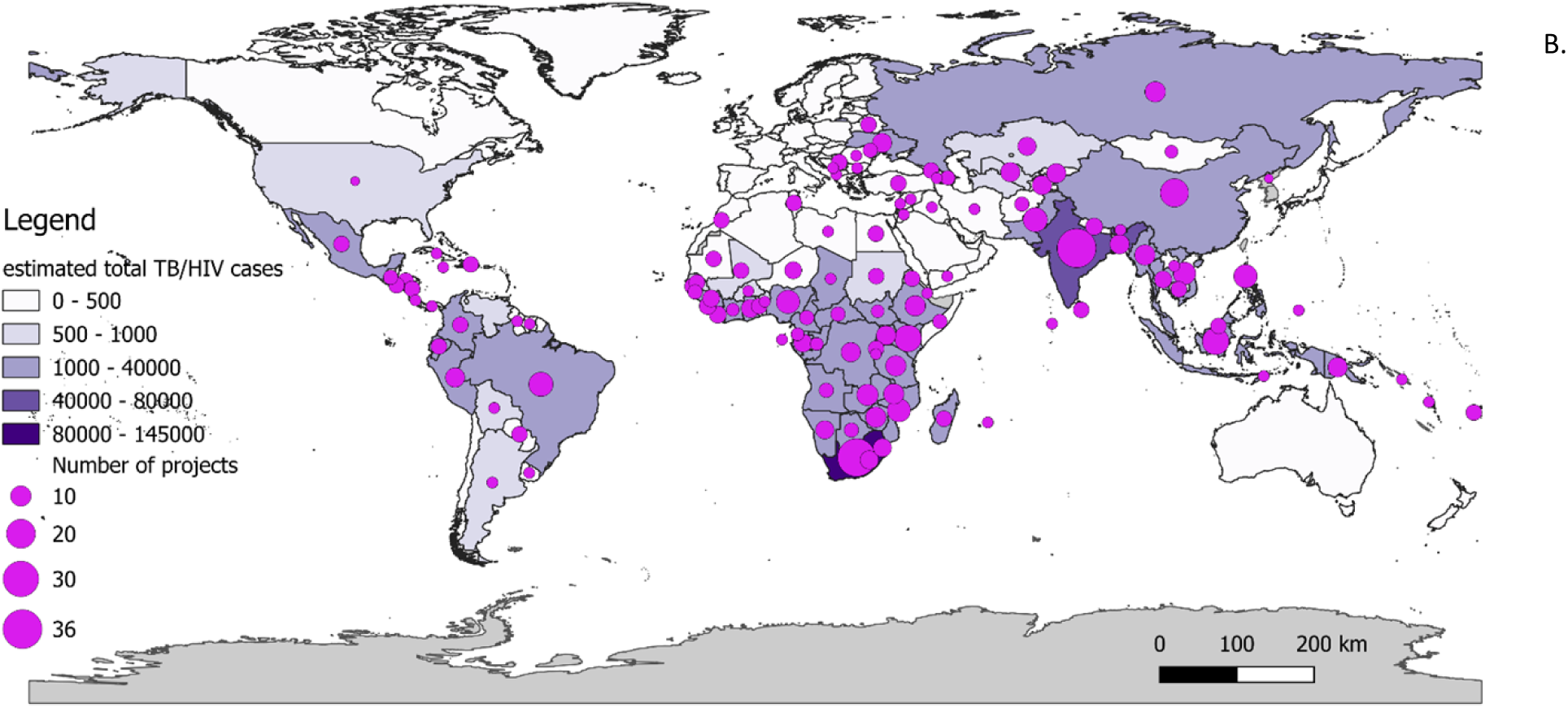
A. World map with the number of adult and adolescent TB vaccine preparedness projects/initiatives and the HIV prevalence in the estimated TB incident cases (%) in 2023 and B. total estimated TB/HIV cases (25).

**Figure S.2.**
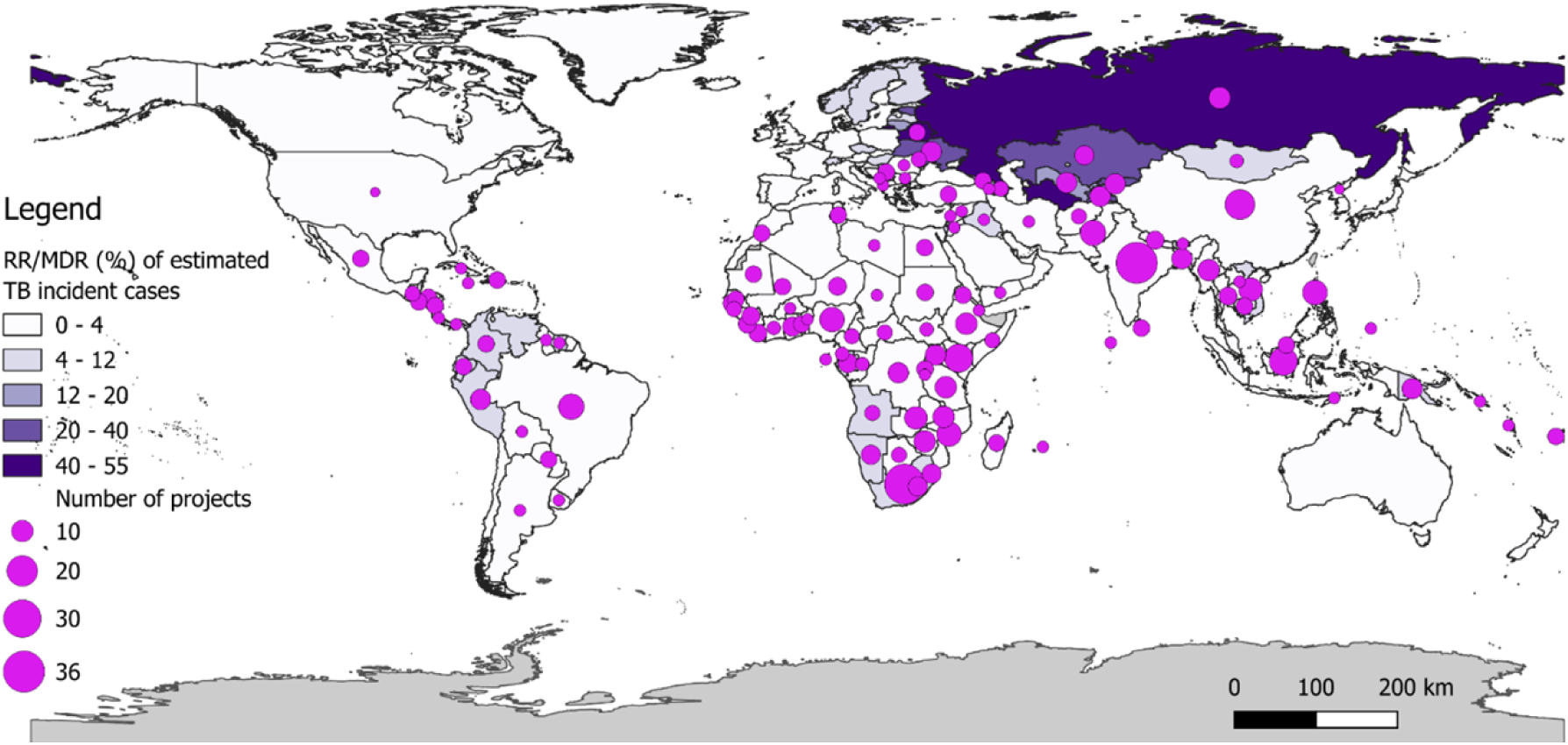
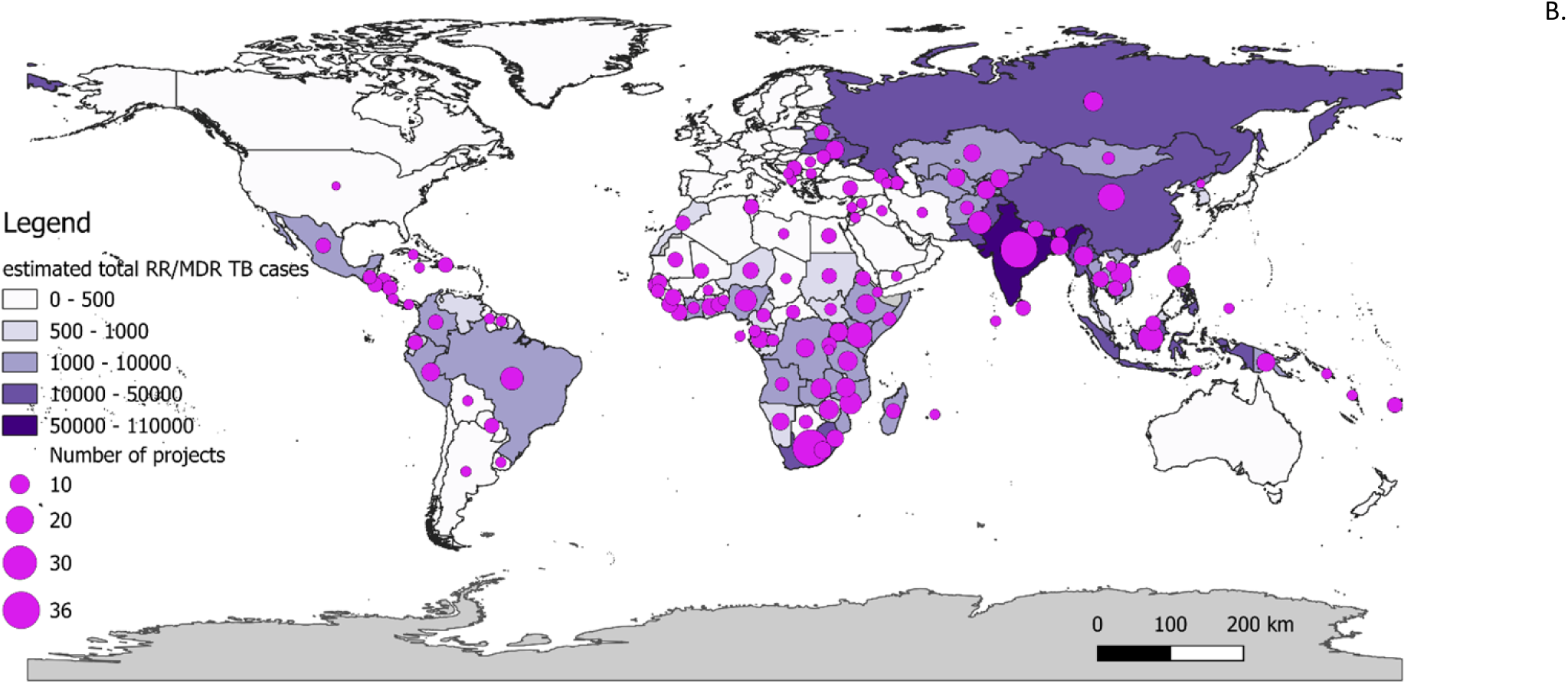
A. World map with the number of adult and adolescent TB vaccine preparedness projects/initiatives and the RR/MDR cases of the estimated TB incident cases (%) in 2023, and B. total estimated RR/MDR cases (25).

**Table S.1.**
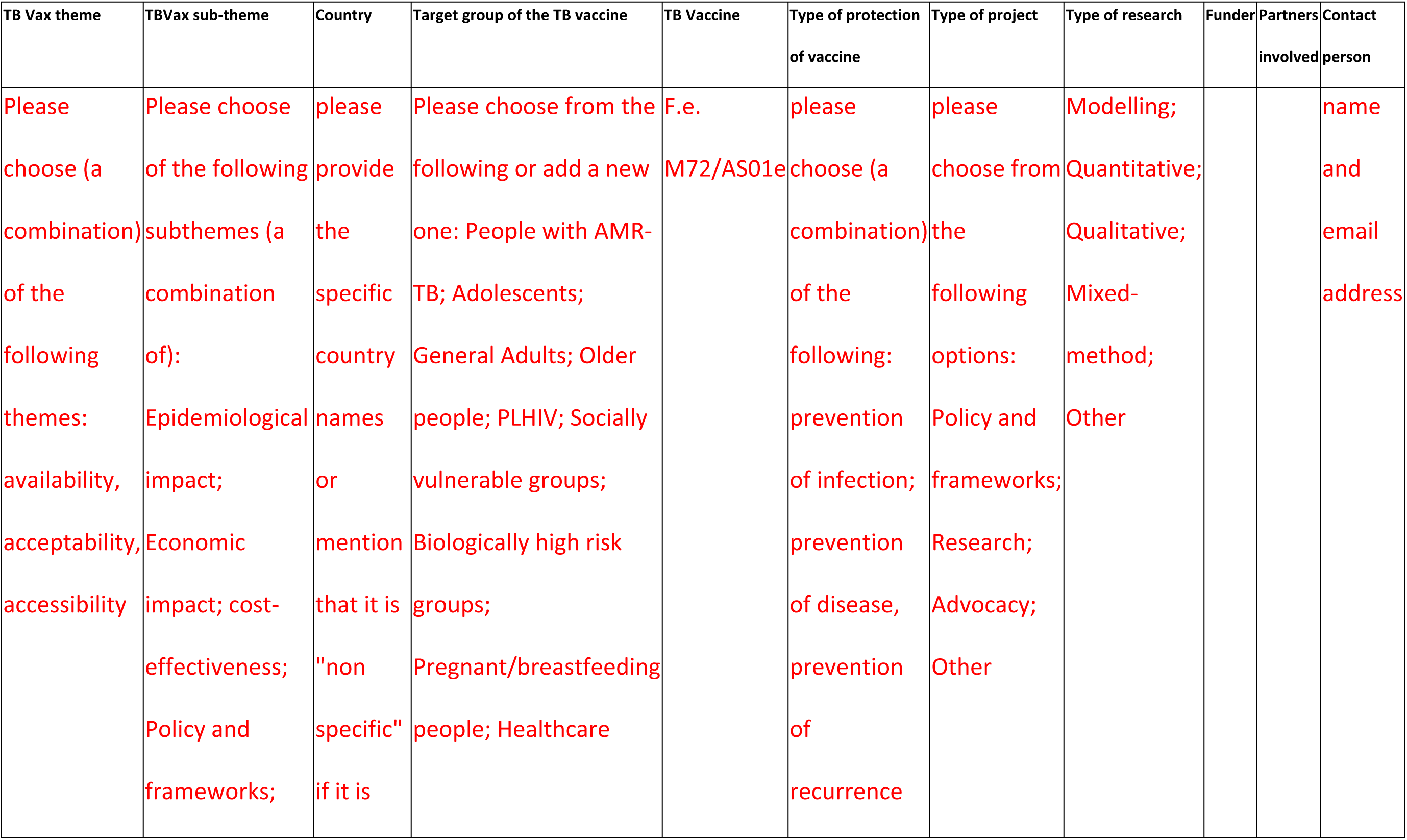
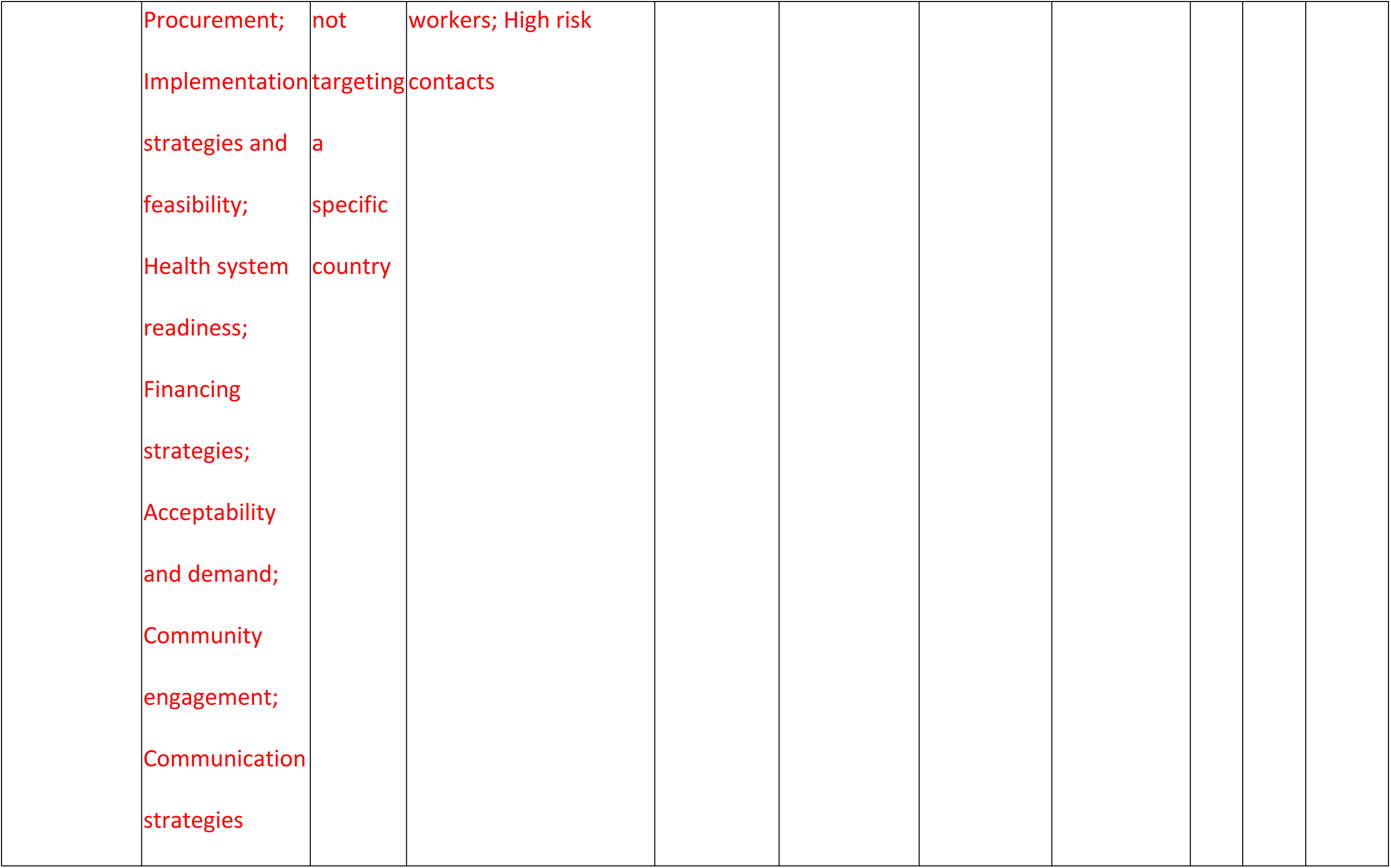
. Project data indicators collected for the novel adult and adolescent TB vaccine preparedness repository.

**Table S.2.**
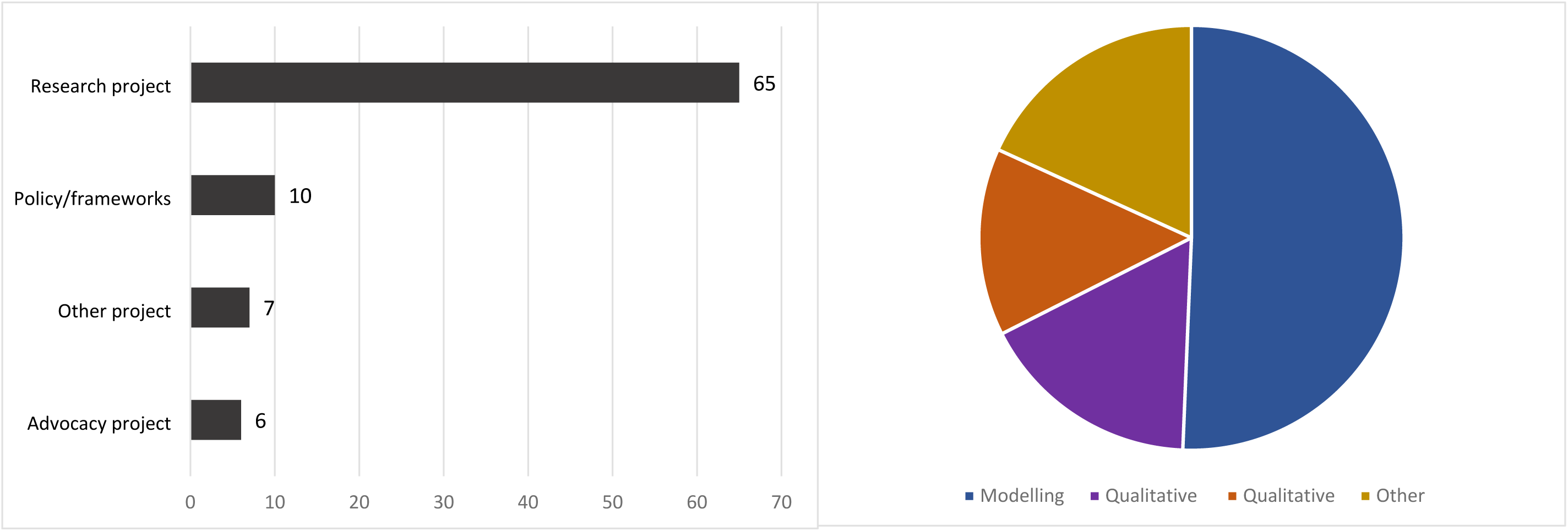
Left: Overview of the types of projects and initiatives (research, policy & frameworks, advocacy, and other project) included in the TB Vaccine Preparedness Repository. Right: Overview of the types of research (modelling, qualitative, quantitative, other) included in the TB Vaccine preparedness repository.

**Table S.3.**
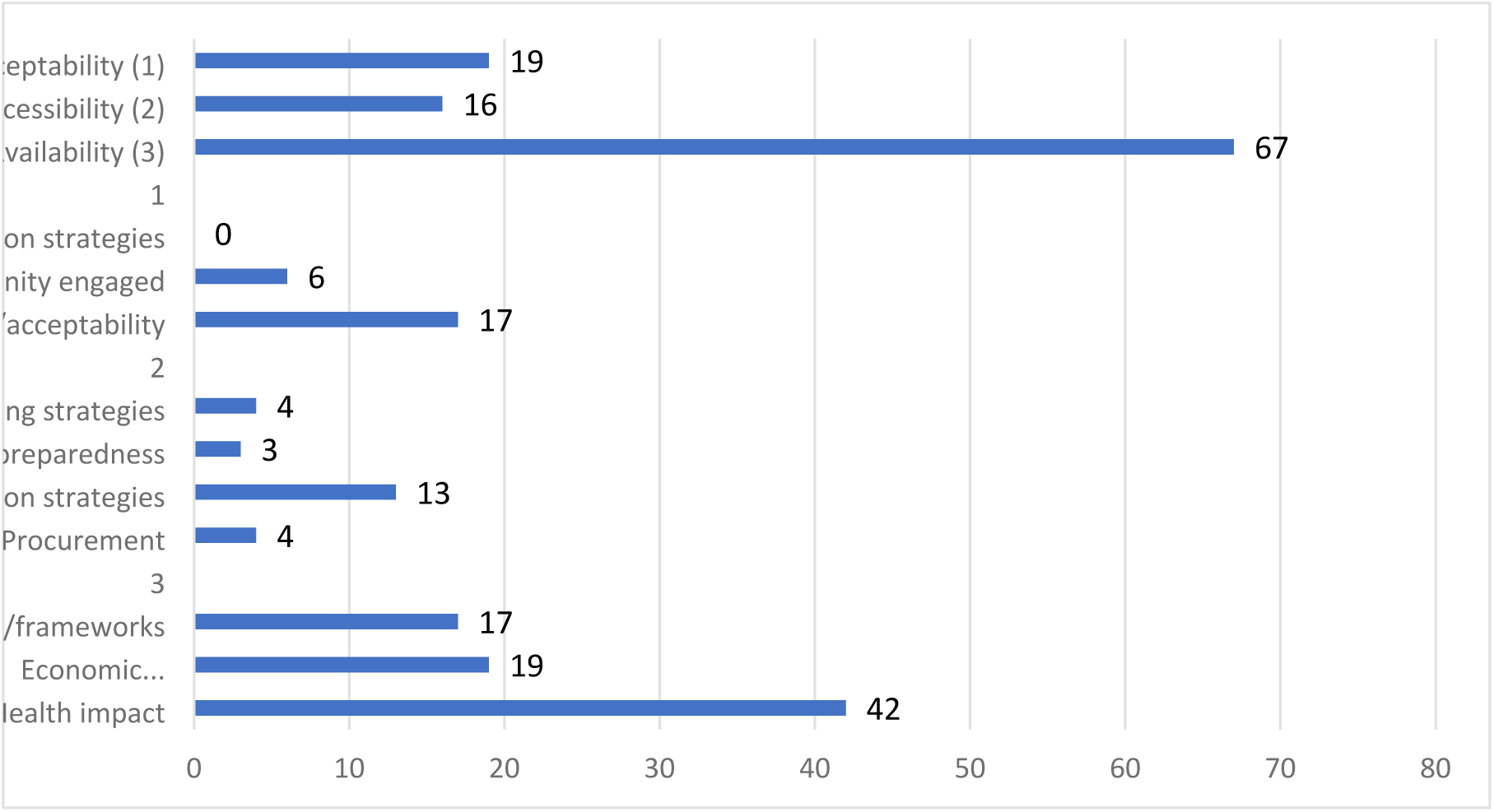
Overview of the adult and adolescent TB vaccine preparedness projects along the WHO’s global framework to prepare for country introduction of new TB vaccines for adults and adolescents that are included in the TB Vaccine Preparedness Repository.

**Table S.6.**
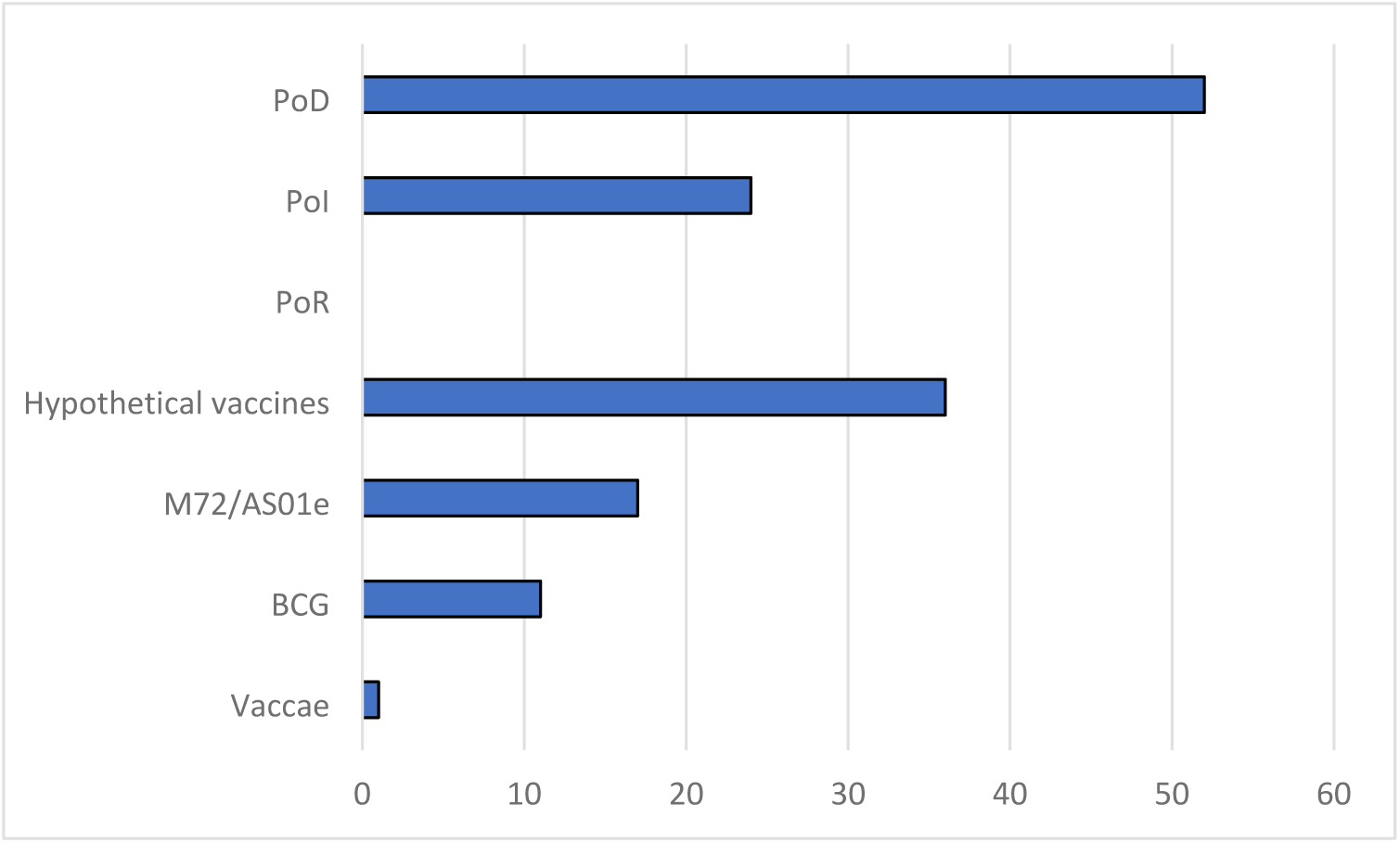
Overview of the number of adult and adolescent TB vaccine preparedness projects according to type of protection (PoD, PoI, PoR) and type of vaccine (BCG revaccination, M72/AS01e, Vaccae) in the TB vaccine preparedness repository.

